# Novel Coronavirus Pandemic Impacts Children and Adolescents’ Psychological Well-Being in Heavily Hit Chinese Provinces

**DOI:** 10.1101/2021.08.27.21262700

**Authors:** Jing Ma, Jun Ding, Jiawen Hu, Kai Wang, Shuaijun Xiao, Ting Luo, Shuxiang Yu, Chuntao Liu, Yunxuan Xu, Yingxian Liu, Changhong Wang, Suqin Guo, Xiaohua Yang, Haidong Song, Yaoguo Geng, Yu Jin, Huayun Chen, Chunyu Liu

**Affiliations:** Department of Child and Adolescent Psychiatry, Brain Hospital of Hunan Province (School of Clinical Medicine, Hunan University of Chinese Medicine), Changsha410007, Hunan, China; Department of Social Work, Shenzhen Mental Health Center, Shenzhen, Guangdong, China; Futian hospital for prevention and treatment of chronic disease, Shenzhen, Guangdong, China; Xiangyifurong Middle School of Changsha, Changsha, Hunan, China; The Second Affiliated Hospital of Xinxiang Medical University, Xinxiang, Henan, China; Changsha Changjun Bilingual Experimental Middle School, Changsha, Hunan, China; Mental Health Center Zhejiang University school of Medicine (Hangzhou Seventh People’s Hospital), Hangzhou310013, Zhejiang, China; School of Education, Zhengzhou University, Zhengzhou 45001, Henan, China; School of Public Health, Sun Yat-sen University, Guangzhou, Guangdong, China; Division of Epidemiology and Biostatistics, School of Public Health, University of Illinois at Chicago, Chicago, IL 60612; Department of Psychiatry, SUNY Upstate Medical University, New York, United States

**Keywords:** Novel coronavirus, psychological state, child and adolescent, anxious symptoms, depressive symptoms, compulsive symptoms

## Abstract

In light of the novel coronavirus’s (COVID-19’s) threat to public health worldwide, we sought to elucidate COVID-19’s impacts on the mental health of children and adolescents in China. Through online self-report questionnaires, we aimed to discover the psychological effects of the pandemic and its associated risk factors for developing mental health symptoms in young people.

We disseminated a mental health survey through online social media, WeChat, and QQ in the five Chinese provinces with the most confirmed cases of COVID-19 during the late stage of the country-wide lockdown. We used a self-made questionnaire that queried children and adolescents aged 6 to 18 on demographic information, psychological status, and other lifestyle and COVID-related variables.

A total of 17,740 children and adolescents with valid survey data participated in the study. 10,022 (56.5%), 11,611 (65.5%), 10,697 (60.3%), 6,868 (38.7%), and 6,225 (35.1%) participants presented, respectively, more depressive, anxious, compulsive, inattentive, and sleep-related problems compared to before the outbreak of COVID-19. High school students reported a greater change in depression and anxiety than did middle school and primary school students. Despite the fact that very few children (0.1%) or their family members (0.1%) contracted the virus in this study, the psychological impact of the pandemic was clearly profound. Fathers’ anxiety appeared to have the strongest influence on a children’s psychological symptoms, explaining about 33% of variation in the child’s overall symptoms. Other factors only explained less than 2% of the variance in symptoms once parents’ anxiety was accounted for.

The spread of COVID-19 significantly influenced the psychological state of children and adolescents. It is clear that children and adolescents, particularly older adolescents, need mental health support during the pandemic. The risk factors we uncovered suggest that reducing fathers’ anxiety is particularly critical to addressing young people’s mental health disorders in this time.

## Background

The novel coronavirus (COVID-19) is highly contagious. On August 23, 2020, there were 84,967 confirmed cases of COVID-19 infections in China and 23,057,288 worldwide[1,2]. Thus far, 4,634 people in China and 800,906 in total have died as a result of the virus, and children and adolescents are among the people that it continues to kill [3].

In addition to individuals’ physical health, COVID-19 has taken a toll on people’s psychological health [4-6]. The pandemic has triggered depressive and anxious symptoms in many individuals [6-12] and may interfere with children’s sleeping and attention [7,12-14]. The psychological influence of COVID-19 on children and adolescents may differ from that on adults due to differences in maturity and psychological development [15].

A minority of the studies examining the influence of the COVID-19 outbreak on psychological symptoms has focused on children’s and adolescents’ psychology. The research that has been done has found that children’s psychological state during the outbreak correlates with several factors, including age [16], mandatory quarantine status [17], mother’s occupation [16], whether a family member has been infected with COVID-19 [18], and whether the participant is a high school student [19,20]. However, no research to date has reported whether children’s symptoms of anxiety and depression during the COVID infection bear a relationship to exercise intensity, history of mental disorders, or parents’ anxiety levels about the pandemic.

Based on the research on and clinical reports of children and adolescents with psychological disturbances that have been done thus far during COVID-19, we hypothesize that: (1) the pandemic has caused many young people to experience more depressive and anxious symptoms, as well as a decrease in concentration and and an increase in sleeplessness compared to before the outbreak; (2) during the pandemic, middle school students have experienced more of the aforementioned symptoms than primary school students; (3) mandatory quarantine, children’s perception of high parental anxiety about COVID-19, low intensity of exercise, parental and own infection status, and having parents who work in a medical field are positively associated with anxious and depressive symptoms in children. We performed a large online survey to collect data to test these hypotheses.

## Methods

### Participants

Students aged six to eighteen were the main subjects of the study. They came from the five Chinese provinces that were the hardest hit by COVID-19: Hunan, Hubei, Guangdong, Zhejiang, and Henan.

### Ethics approval

This study has been approved by ‘Ethics Committee of Shenzhen Futian District Chronic Disease Prevention and Treatment Hospital’. The IRB waived the need for the parents’ consent, since it is online anonymous survey. No privacy-related information, like name, address, etc. will be collected.

### Measures

An online questionnaire (www.wjx.cn) was used to create and publish the self-made questionnaire. Social media, WeChat and QQ were used to disseminate the questionnaires, which participants completed between February 19th and March 5th. During this period, most parts of China were still in complete lockdown, which started on January 23^rd^ and ended around late April.

Questions about mental symptoms were written in language that children could understand. When the children had difficulties reading questions or responding, their parents were permitted to help them.

### Questionnaire

The questionnaire contained two parts, one of which included demographic and lifestyle information, and the other of which asked about psychological status of self and parents.

Demographic and lifestyle information included questions about age, place of residence, mental disorder history, whether or not family members had been infected, quarantine status, whether partcipants had had close contact with an infected person, parents’ profession (medical staff member or not, fever clinic staff member or not), parents’ risk of contracting COVID-19, as well as duration and intensity of participant exercise.

The section on psychological status included questions about whether the participant’s parents were more anxious than before the outbreak of COVID-19 in the child’s opinion, as well as whether the children felt more anxious, depressive, compulsive, irritable, and unable to focus and sleep compared to before the outbreak. Children were asked about three anxiety symptoms: inability or difficulty relaxing muscles; excessive worries; and nervousness, fidgeting, and restlessness. Depressive symptoms included four symptoms: low mood; diminished interest; helplessness; and irritability. Compulsive symptoms include compulsive hand-washing and compulsive thinking. Scoring options for the second part of the survey ranged from zero to ten, with a zero indicating the child’s experience of the symptoms were the same as before the outbreak and a ten indicating their symptoms were the worst they had ever experienced.

### Quality Control

The survey used a web-based questionnaire, and only one response could be given per IP address. To be included in the study, participants needed to: (1) be between the ages of six and eighteen; (2) have completed the questionnaire in more than one minute; (3) resided in one of the five following provinces: Hubei, Hunan, Henan, Zhejiang, or Guangzhou. All other participants were excluded. Participants were also excluded if their responses did not match the questions. We initially collected 20,677 questionnaires; after some responses were eliminated according to the aforementioned exclusion criteria, 17,740 valid questionnaires remained.

### Statistical analyses

Data were analyzed using SPSS version 25.0. Descriptive statistics were obtained first. Logistic regression analyses were then used to compare the risk of anxious symptoms, depressive symptoms, compulsive symptoms, inattentive symptoms, and sleep problems between primary, middle, and high school students, where odds ratios were calculated for high school students and middle school students with respect to primary school students. To examine factors influencing children’s psychological state during the COVID-19 outbreak, a principal component analysis of the outcomes, including anxious, depressive, and compulsive symptoms, was performed. The first principal component captured 65% of variation in outcomes. The first principal component was used as the major dependent variable in the subsequent multivariate linear regression analysis of the risk factors. The independent variables, (e.g. mental disorder, mandatory isolation, exercise intensity), were examined for their effect on children’s psychological state during the COVID-19 outbreak. Supplemental analyses of the residual scores after the first principal component had been removed from the outcomes were also performed using the factor analysis and multivariate regression analyses. In all the analyses, we use a p-value of 5% as the cutoff in analyzing significance of risk factors.

## Results

### 1. Characteristics of the study population

A total of 17,740 children and adolescents participated in the study. Table 1 lists summary statistics of the study sample. There were 8,182 (46.1%) primary school students (age: 6 - 11), 6,543 (36.9%) middle school students (age: 12-14) and 3,015 (17.0%) high school students (age: 15-18). The regional distribution of the participants was as follows: Hubei--305 (1.7%), Zhejiang--1,314 (7.4%), Henan--3,541 (20.0%), Hunan--4,779 (26.9%), Guangdong--7,801 (44.0%). Among the participants, 13 (0.1%) were knowingly infected by COVID-19,12 (0.1%) had knowledge of an infected family members, and only 1 (0.01%) out of the 12 infected had a family member who passed away from COVID-19. There were 15,444 (87%) respondents under voluntary home quarantine, and 1,201 (6.8%) individuals were under mandated quarantined by order of the government, hospitals, and communities.

**Table 1.** The incidence and mean score for different psychological symptoms (N=17,740)

| Symptoms | Symptoms<br>Worsened<br>during<br>COVID? | n (%) | Mean (95% CI) |
| --- | --- | --- | --- |
| Muscle Tension | Yes | 8742 (49.3%) | 1.63 (1.59-1.66) |
|  | No | 8998 (50.7%) |  |
| Excessive worries | Yes | 10014 (56.4%) | 1.91 (1.88-1.95) |
|  | No | 7726 (43.6%) |  |
| Fidgeting | Yes | 9096 (51.1%) | 1.74 (1.71-1.78) |
|  | No | 8671 (48.9%) |  |
| Low mood | yes | 8168 (46%) | 1.55 (1.52-1.58) |
|  | no | 9572 (54%) |  |
| diminished interest | yes | 7066 (39.8%) | 1.39 (1.35-1.42) |
|  | no | 10674 (60.2%) |  |
| Despair | yes | 5589 (31.5%) | 1.04 (1.01-1.07) |
|  | no | 12151 (68.5%) |  |
| Excessive hand washing | yes | 9357 (52.7%) | 1.66 (1.62-1.69) |
|  | no | 8383 (47.3%) |  |
| Compulsive thoughts | yes | 8506 (47.9%) | 1.35 (1.32-1.38) |
|  | no | 9234 (52.1%) |  |
| Quick to lose temper | yes | 6420 (36.2%) | 1.15 (1.12-1.18) |
|  | no | 11320 (63.8%) |  |
| Inattention | yes | 6868 (38.7%) | 1.25 (1.22-1.28) |
|  | no | 10872 (61.3%) |  |
| Sleep problems | yes | 6225 (35.1%) | 1.29 (1.26-1.32) |
|  | no | 11515 (64.9%) |  |

Table 2 displays the summary statistics of the impact of COVID-19 on children’s psychological symptoms. The table shows that, among the participants, 10,022 (56.5%), 11,611 (65.5%), 10,697 (60.3%), 6,868 (38.7%), and 6,225 (35.1%), respectively, presented more depressive, anxious, compulsive, inattentive, and sleep problems than before the outbreak of COVID-19.

**Table 2.** The incidence of psychological symptoms among high school, middle school, and primary school students (before versus after the outbreak) (N=17,740)

| **More mental**<br>**Symptoms than before** | **Primary school**<br>**students(n=8182)** | **Middle school**<br>**students(n=6543)** | **High school**<br>**students(n=3015)** |
| --- | --- | --- | --- |
| Depressive symptoms | 4552 (55.6%) | 3670 (56.0%) | 1800 (59.7%) |
| Anxious symptoms | 5252 (64.2%) | 4284 (65.5%) | 2075 (68.8%) |
| Compulsive symptoms | 4933 (60.3%) | 3950 (60.4%) | 1814 (60.2%) |
| Inattention | 3113 (38.0%) | 2452 (37.5%) | 1303 (43.2%) |
| Sleep problems | 2871 (35.0%) | 2207 (33.7%) | 1147 (38.0%) |

### 2. The severity of psychological symptoms increases with age: comparison of the impact on children from primary, middle and high schools

Table 3 shows the results of the logistic regression analyses measuring the association between psychological symptoms and school level. As depicted in Table 3, there was no difference between primary and middle school students on anxiety and depression, or on problems with sleep and attention (p>0.05),but there were differences between primary and high school students (p<0.05). High school students reported more anxiety, depression, attention deficit, and sleep problems than primary school students (OR high school students>1, OR primary school students=1). There were no differences between compulsive symptoms in high school students, middle school students, and primary school students (p>0.05).

**Table 3.** Comparison between school grade levels and psychological symptoms.

| **Variables** | **primary school** | **middle school** | **high school** |
| --- | --- | --- | --- |

|  |  | students | students | students |
| --- | --- | --- | --- | --- |
| Depressive | OR | 1 | 1.019 | 1.181 |
| symptoms | 95% CI | - | 0.954-1.088 | 1.085-1.286 |
|  | P | - | 0.58 | <0.001 |
| Anxious | OR | 1 | 1.058 | 1.231 |
| symptoms | 95% CI | - | 0.988-1.133 | 1.126-1.347 |
|  | P | - | 0.105 | <0.001 |
| Compulsive | OR | 1 | 1.003 | 0.995 |
| symptoms | 95% CI | - | 0.939-1.072 | 0.913-1.083 |
|  | P | - | 0.922 | 0.905 |
| Inattention | OR | 1 | 0.976 | 1.239 |
|  | 95% CI | - | 0.913-1.044 | 1.139-1.349 |
|  | P | - | 0.477 | <0.001 |
| Sleep problems | OR | 1 | 0.942 | 1.136 |
|  | 95% CI | - | 0.879-1.008 | 1.042-1.238 |
|  | P | - | 0.085 | <0.001 |

### 3. Factors influencing children’s psychological state during the COVID-19 outbreak

Since depressive symptoms, anxious symptoms, and compulsive symptoms measure some similar psychological features, we first performed a principal component analysis. The results are displayed in Tables 4 and 5.

**Table 4.**
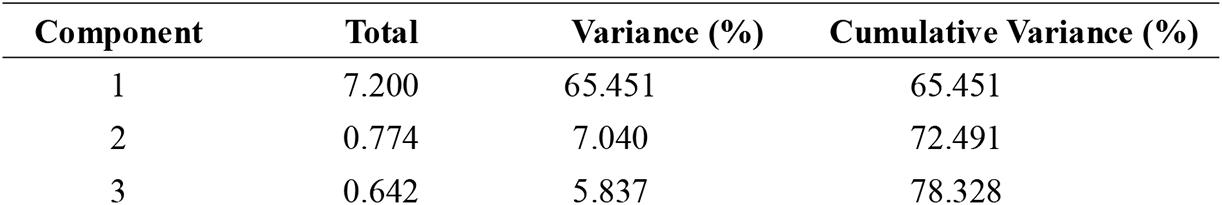
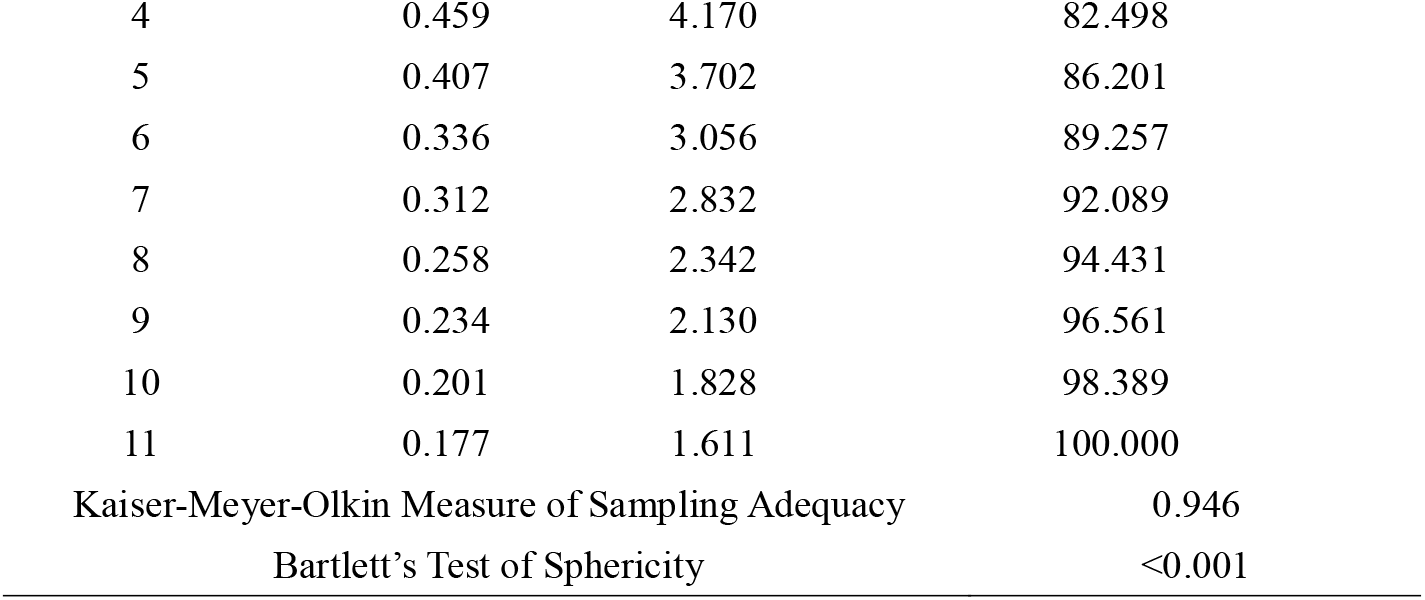
Characteristic values of input data, cumulative variance contribution rate and KMO and Bartlett’s Test.

| Component | Total | Variance (%) | Cumulative Variance (%) |
| --- | --- | --- | --- |
| 1 | 7.200 | 65.451 | 65.451 |
| 2 | 0.774 | 7.040 | 72.491 |
| 3 | 0.642 | 5.837 | 78.328 |
| 4 | 0.459 | 4.170 | 82.498 |
| 5 | 0.407 | 3.702 | 86.201 |
| 6 | 0.336 | 3.056 | 89.257 |
| 7 | 0.312 | 2.832 | 92.089 |
| 8 | 0.258 | 2.342 | 94.431 |
| 9 | 0.234 | 2.130 | 96.561 |
| 10 | 0.201 | 1.828 | 98.389 |
| 11 | 0.177 | 1.611 | 100.000 |
| Kaiser-Meyer-Olkin Measure of Sampling Adequacy |  |  | 0.946 |
| Bartlett's Test of Sphericity |  |  | <0.001 |

**Table 5.** Principal component analysis results.

| Variable | Variance Explained by Component 1 |
| --- | --- |
| Low mood | 0.862 |
| Excessive worries | 0.858 |
| Fidgeting | 0.824 |
| diminished interest | 0.823 |
| Despair | 0.822 |
| Compulsive thoughts | 0.820 |
| Inattention | 0.814 |
| Easy to lose temper | 0.803 |
| Muscle Tension | 0.800 |
| Excessive hand washing | 0.736 |
| Sleep problems | 0.726 |

The first principal component explained 65.45% of the total variation of all the outcomes. In comparison, the second principal component only accounted for 7% of the total variation (Table 4). The first principal component explained an approximately equal proportion of variance for each of the standardized outcome variables (see Table 5). To make the results more interpretable, total standardized symptom score was used instead of the first principal component in the following regression analyses.

Table 6 displays the results of the linear regression, where the total score is the outcome and the covariates were selected using the stepwise variable selection approach. Interestingly, father’s anxiety about the virus contributed substantially to a child’s psychological symptoms (R^2^=0.33) while, in comparison, mother’s anxiety about the virus only had a small additional effect on the child’s psychological symptoms (R^2^=0.02) once the father’s anxiety was accounted for. Although the father’s anxiety was correlated with the mother’s anxiety, it appears that the father’s anxiety had a stronger influence on a child’s psychological symptoms than the mother’s anxiety.

**Table 6.**
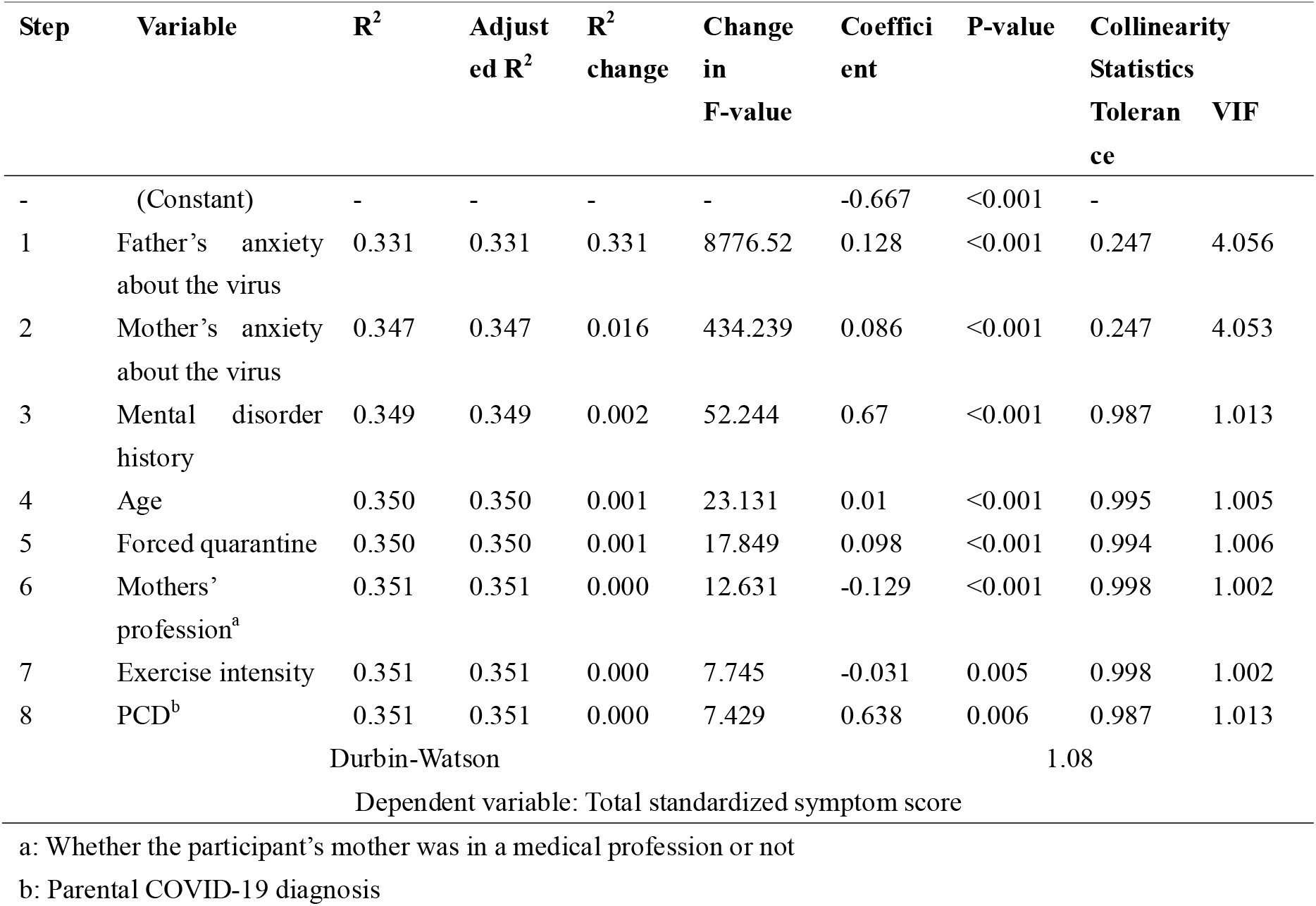
Analysis of factors influencing total score.

| Step | Variable | R<sup>2</sup> | Adjusted R<sup>2</sup> | R<sup>2</sup> change | Change in F-value | Coefficient | P-value | Collinearity Statistics Tolerance | VIF |
| --- | --- | --- | --- | --- | --- | --- | --- | --- | --- |
| - | (Constant) | - | - | - | - | -0.667 | <0.001 | - |  |
| 1 | Father's anxiety about the virus | 0.331 | 0.331 | 0.331 | 8776.52 | 0.128 | <0.001 | 0.247 | 4.056 |
| 2 | Mother's anxiety about the virus | 0.347 | 0.347 | 0.016 | 434.239 | 0.086 | <0.001 | 0.247 | 4.053 |
| 3 | Mental disorder history | 0.349 | 0.349 | 0.002 | 52.244 | 0.67 | <0.001 | 0.987 | 1.013 |
| 4 | Age | 0.350 | 0.350 | 0.001 | 23.131 | 0.01 | <0.001 | 0.995 | 1.005 |
| 5 | Forced quarantine | 0.350 | 0.350 | 0.001 | 17.849 | 0.098 | <0.001 | 0.994 | 1.006 |
| 6 | Mothers' profession<sup>a</sup> | 0.351 | 0.351 | 0.000 | 12.631 | -0.129 | <0.001 | 0.998 | 1.002 |
| 7 | Exercise intensity | 0.351 | 0.351 | 0.000 | 7.745 | -0.031 | 0.005 | 0.998 | 1.002 |
| 8 | PCD<sup>b</sup> | 0.351 | 0.351 | 0.000 | 7.429 | 0.638 | 0.006 | 0.987 | 1.013 |
| Durbin-Watson |  |  |  |  |  |  | 1.08 |  |  |
| Dependent variable: Total standardized symptom score |  |  |  |  |  |  |  |  |  |

a: Whether the participant's mother was in a medical profession or not
b: Parental COVID-19 diagnosis

Other factors that had significant effects on the child’s psychological symptoms included age, history of mental disorders, mandatory quarantine status, whether their mother worked as a medical staff employee, exercise intensity, and whether a parent had been infected with COVID-19 to the child’s knowledge. However, these factors explained less than 2% of the variation in the child’s symptoms once parents’ anxiety was accounted for. Given the huge sample size in this study, the importance of these significant results that add little interpretive power (R^2^) can be limited.

To examine if there were any important risk factors that explained the remaining variance in the child’s symptoms other than the total score, we examined the residual scores obtained by projecting the outcomes to the orthogonal complement space of the total score. Factor analysis was performed on the residual scores, and regression analyses were then performed using the identified factors as outcomes and the risk factors as covariates. The results of these analyses suggest that the covariates do not explain a major proportion of the residuals. The results of these additional analyses are provided in the supplemental materials.

## Discussion

In this study, we explored levels of psychological distress since the COVID-19 pandemic in 17,740 children and adolescents. The six-to-eighteen-year-old students, all of whom resided in the five Chinese provinces with the highest number of reported cases, responded to questionnaires regarding whether their depressive, anxious, compulsive, inattentive, and sleep-related issues had heightened since the beginning of the pandemic.

More than half of the participants experienced more anxious and depressive symptoms than they had before the outbreak, and about one-third of the participants experienced more sleeplessness and inattentive symptoms. Though reports of compulsion were approximately equal across primary school, middle school, and high school students, we found that high school students reported significantly more worsening of anxiety, depression, inattention, and trouble with sleep than did primary school students. Previous research has found that grade level is positively correlated with depression and anxiety sympoms [19,20]. Additionally, because our study asked participants about their experience of symptoms as compared to their normal state, our results suggests that, when compared to younger children, adolescents’ mood and anxiety are more susceptible to change as a result of the current pandemic conditions (as opposed to simply having a higher baseline rate of depression and anxiety).

Since COVID-19, the rates of anxiety and depression in Chinese adults have been reported at 6.33%-32.1% and 16.5%-17.17%, respectively [15,21], and 57.1% have reported poor sleep [22]. 43.7% of Chinese high school student have been experiencing depressive symptoms, and 37.4% have been experiencing anxiety symptoms [19]. The reported prevalence of depressive symptoms among children and adolescents in China during the pandemic have been somewhat lower than for adolescents, at 22.28% [16]. In contrast, during non-epidemic periods, the prevalence of depression and anxiety in China was 6.8% and 7.6% in the general population [23], and ∼3.5%-44.0% for depression and ∼20.31%-26.70% for anxiety in children and adolescents [23]. Because only a small proportion of the children in this study had confirmed COVID-19 diagnoses, it appears their psychological symptoms were caused primarily by the psychological distress around the conditions coinciding with the pandemic, not the health effects of the virus itself.

When we examined the relationship between independent variables and degree of psychological distress, we found that the children’s perception of their fathers’ anxiety about COVID-19 explained a sizable proportion (∼33%) of variance in reported changes in psychological distress, while perception of mothers’ anxiety explained only a small proportion of variance after father’s anxiety was accounted for. This relationship between children’s self-rated total scores of psychological symptoms and the belief that their parents were anxious about the pandemic has not been reported before, nor was this trend reported during SARS. Though we cannot draw a definitive directional or causal conclusion between the two factors or know whether the children’s perceptions were accurate, it is possible that father’s anxiety about COVID-19 in children’s eyes had more of an impact on children’s depressive and anxious symptoms than that of their mothers because mothers are usually more sensitive and their emotions change more frequently than fathers’ emotions [24,25], which could lead children to become desensitized to their mothers’ emotions. In contrast, fathers usually do not visibly display anxiety [24,25], which could make children more likely to sense it in their fathers when it is displayed. The fathers’ stronger impact on children’s emotion was reported before [26,27]. Previous research has addressed the point that when times are tough, parental support can ease children’s stress and anxiety, but amidst the outbreak of COVID-19, parents have also been feeling a lot of anxiety due to the risk of infection and financial stress. This likely leaves parents with fewer emotional resources with which to support their children. As a result, children’s perception of their parents’ anxiety could in turn increase their own anxiety [28].

Age of participant, history of mental disorders, mandatory quarantine status, whether the participant’s mother worked as a medical staff member, exercise intensity, and parent infection status had significant effects on the child’s psychological symptoms but, in total, still explained less than 2% of the variation in symptoms once parents’ anxiety was accounted for. Some of these findings were consistent with previous studies in adults [14,15,6,5,29] and children (specifically with regard to effects of age [16], quarantine status [17], [21,30], mother’s profession [16], and having a family member infected with COVID-19 [18]). Some research has found that participants with a history of mental disorders were more vulnerable to stressful events and likely to have more emotional problems than the general population during the pandemic, though tehse studies used relatively qualitative methods [31,32]; many previous studies have suggested that lower exercise intensity is related to more symptoms of depression and anxiety in adolescents, though these studies were conducted outside the context of the COVID-19 pandemic [33-36].

Though the significant independent variables, aside from paternal anxiety, only explain a small proportion of variance in children’s psychological wellbeing after parental anxiety is accounted for, they reveal interesting connections between mental health of youth and life factors in an unusal historical moment.

The findings above have important clinical implications. For example, we found evidence that children and adolescents, especially those with a history of mental disorders, may need more psychological attention and care during a public health emergency such as the one that the world is currently experiencing. Additionally, management of parents’ (especially fathers’) own anxiety may help their children’s mental wellbeing.

One limitation of the current study is the fact that all participants filled in the questionnaire online without supervision, though rigorous quality control was implemented. Additionally, the sample size collected in Hubei, the hardest-hit area, is the smallest in this study. The respondents in this area may not represent subjects in the area well in comparison to participants in the other provinces.

In conclusion, the COVID-19 outbreak has influenced Chinese children’s and adolescents’ psychological states significantly. More than half of the participants experienced more anxious and depressive symptoms than before, and about one-third of the participants had more sleepless and inattentive symptoms. With respect to their baseline states of mood and anxiety, adolescents were more anxious than younger children, and high school students were more depressive and anxious than middle school and primary school students. Father’s perceived anxiety about the virus had a substantially stronger influence on a child’s psychological symptoms than mother’s anxiety did. Other factors, including age, history of mental disorders, mandatory quarantine status, whether the participant’s mothers worked as a medical staff member, exercise intensity, and whether the child’s parents had been infected with COVID-19, were also found to have significant effects on the child’s psychological symptoms. Stil, when added together, they explained less than 2% of variances in the child’s symptoms once parents’ anxiety was accounted for.

The crucial takeaways from our study suggest that, though both have been effected, the COVID-19 pandemic has had a heavier impact on adolescent mental health than it has on mental health of younger individuals and that paternal anxiety about the pandemic is heavily related to children and adolescents’ psychological states. Future research should further investigate other mechanisms that may be moderating parental and children’s anxiety, as well as whether paternal anxiety precedes the child’s anxiety about COVID-19 or vice versa. Then, researchers will better be able to work towards mitigating the effects of mental health issues in high-anxiety families within the context of large crises such as the current pandemic.

## Data Availability

The datasets used and/or analyzed during the current study are available from the corresponding author on reasonable request.

## Declaration

### Ethics approval

This study has been approved by ‘Ethics Committee of Shenzhen Futian District Chronic Disease Prevention and Treatment Hospital’. And it has therefore been performed in accordance with the ethical standards laid down in the 1964 Declaration of Helsinki and its later amendments. The specific national laws have been observed, too.

### Consent to participate

Not Applicable. This is not a clinical trial, and we did not investigate the participants’ names and other private information when we conducted the questionnaire.

### Consent of publication

All the authors signed a consent form for publication

## Authors’ contributions

Jing Ma, Jun Ding and Jiawen Hu contributed equally to the work as co-first authors.

Huayun Chen and Chunyu Liu contributed equally to the work as co-corresponding authors. Jing Ma had full access to all the data in the study and takes responsibility for the integrity of the data and the accuracy of the data analysis. Concept and design: Jing Ma, Chunyu Liu, Jun Ding. Acquisition, analysis, or interpretation of data: All authors. Drafting of the manuscript: Jing Ma, Jiawen Hu, Chunyu Liu, Huayun Chen, Jun Ding. Critical revision of the manuscript for important intellectual content: All authors. Statistical analysis: Huayun Chen, Jiawen Hu, Jing Ma, Shuaijun Xiao, Ting Luo. Obtained funding: Jing Ma, Jun Ding. Administrative, technical, or material support: Kai Wang, Shuxiang Yu, Chuntao Liu, Yunxuan Xu, Yingxian Liu, Changhong Wang, Suqin Guo, Xiaohua Yang, Haidong Song, Yaoguo Geng, Yu Jin

## Acknowledgments

We would like to acknowledge the support offered by the project of Technological Innovation Guidance Plan of Hunan province (a randomized controlled study of the effect of social skills training on Asperger syndrome, 2017SK50314), a project from Hunan Provincial Commission of Health (Project number: B2019045), the Diagnosis and Treatment Enhancement Project of Hunan Provincial Severe Mental Illness (Project number: 2018SK7002), the Autism Center of Hunan Provincial Key Discipline Project, and the Shenzhen Fund for Guangdong Provincial High Level Clinical Key Specialties (Project number: SZGSP013).

