## supplemental material for "Novel Coronavirus Pandemic Impacts Children and Adolescents’ Psychological Well-Being in Heavily Hit Chinese Provinces"

The residuals for the total symptom scores (i.e., the projection of the outcomes to the orthogonal complement space of the total score) were examined. A factor analysis was first performed on the residual scores and regression analyses were then performed using the identified factors as outcomes and the risk factors as covariates.

The average score, C1= (ZX1+ZX2 +ZX3+ZX4+ZX5+ZX6+ZX7+ZX8+ZX9 +ZX10+ZX11)/11, was used in the first regression analysis. Next, we used ZX1 as the outcome and C1 as the covariate, ran a univariate regression, and obtained the residuals of the regression, denoted WX1. We repeated the same regression analysis with ZX1 replaced by ZX2, ZX3, etc. in subsequent regressions to obtain the corresponding residuals, denoted WX2, WX3, etc. We then used the residuals (i.e., WX1, WX2 …, WX11) as the inputs for the factor analysis. In short, we kept the first interpretable component (slightly distinct from the first principal component) fixed and rotated the rest of the components.

The results of the factor analysis and regression analysis of residuals WX1-WX11 are as follows：

**Total Variance Explained**

| **Component** | **Initial Eigenvalues** | | | **Rotation Sums of Squared Loadings** | | |
| --- | --- | --- | --- | --- | --- | --- |
|  | **Total** | **% of Variance** | **Cumulative %** | **Total** | **% of Variance** | **Cumulative %** |
| 1 | 2.213 | 20.115 | 20.115 | 1.959 | 17.811 | 17.811 |
| 2 | 1.818 | 16.530 | 36.645 | 1.670 | 15.184 | 32.995 |
| 3 | 1.278 | 11.621 | 48.266 | 1.443 | 13.120 | 46.115 |
| 4 | 1.074 | 9.764 | 58.030 | 1.311 | 11.915 | 58.030 |
| 5 | 1.013 | 9.213 | 67.243 |  |  |  |
| 6 | .956 | 8.690 | 75.933 |  |  |  |
| 7 | .720 | 6.548 | 82.481 |  |  |  |
| 8 | .686 | 6.232 | 88.713 |  |  |  |
| 9 | .678 | 6.162 | 94.876 |  |  |  |
| 10 | .564 | 5.124 | 100.000 |  |  |  |
| 11 | 9.283E-15 | 8.439E-14 | 100.000 |  |  |  |

| **Rotated Component Matrix^a^** | | | | |
| --- | --- | --- | --- | --- |
|  | Component | | | |
|  | 1 | 2 | 3 | 4 |
| WX Loses temper easily | -.741 | .140 | 0.012 | -.233 |
| WX Muscle tension | .715 | .121 | -.037 | -.321 |
| WX Worries too much | .693 | .113 | .131 | -.392 |
| WX Inattention | -.616 | .175 | -.306 | -.250 |
| WX Excessive hand washing | .065 | -.779 | -.054 | -.008 |
| WX Excessive thinking | -.009 | -.774 | -.063 | -.057 |
| WX Sleep problems | -.140 | .270 | -.693 | .051 |
| WX Low spirits | -.031 | .308 | .667 | .230 |
| WX Fidgeting | .100 | .355 | .630 | -.267 |
| WX Loss of interest | .042 | .289 | -.029 | .651 |
| WX Despair | -.058 | -.094 | .013 | .620 |
| Extraction Method: Principal Component Analysis.  Rotation Method: Varimax with Kaiser Normalization. | | | | |
| a. Rotation converged in 7 iterations. | | | | |

**Table 1 Regression analysis of the first principal component on the covariates**

| **step** | **Variable** | **R2** | **Adjusted R2** | **R2 change** | | **F change** | **B** | **P** | **Tolerance** | **VIF** |
| --- | --- | --- | --- | --- | --- | --- | --- | --- | --- | --- |
| 0 | （Constant） |  |  |  | |  | -0.341 | <0.001 |  |  |
| 1 | Father’s nervousness | 0.051 | 0.051 | 0.051 | | 957.08 | 0.052 | <0.001 | 0.246 | 4.06 |
| 2 | Mother’s nervousness | 0.053 | 0.053 | 0.002 | | 36.922 | 0.032 | <0.001 | 0.246 | 4.059 |
| 3 | Exercise duration | 0.055 | 0.055 | 0.002 | | 32.907 | 0.033 | <0.001 | 0.893 | 1.12 |
| 4 | PD^a^ | 0.056 | 0.056 | 0.001 | | 19.464 | -0.516 | <0.001 | 0.999 | 1.001 |
| 5 | PR^b^ | 0.056 | 0.056 | 0 | | 7.513 | -0.071 | 0.007 | 0.994 | 1.006 |
| 6 | Neighbor status^c^ | 0.057 | 0.056 | 0 | | 4.227 | -0.076 | 0.034 | 0.995 | 1.005 |
| 7 | Exercise intensity | 0.057 | 0.056 | 0 | | 3.859 | 0.028 | 0.049 | 0.894 | 1.119 |
| Durbin-Watson | | | | | 1.71 | | | | | |
| Dependent variable: First Principal Component | | | | | | | | | | |

a: history of physical disorder

b: risk of infection in parents

c: neighbor’s COVID-19 infection status

**Table 2 Regression analysis of the second principal component on the covariates**

| **step** | **Variable** | **R2** | **Adjusted R2** | **R2 change** | | **F change** | **B** | **P** | **Tolerance** | **VIF** |
| --- | --- | --- | --- | --- | --- | --- | --- | --- | --- | --- |
| 0 | (Constant) |  |  |  | |  | 0.06 | 0.148 |  |  |
| 1 | Exercise time | 0.006 | 0.006 | 0.006 | | 112.454 | -0.058 | <0.001 | 0.887 | 1.128 |
| 2 | Age | 0.009 | 0.008 | 0.002 | | 40.004 | 0.016 | <0.001 | 0.988 | 1.012 |
| 3 | Father nervous | 0.01 | 0.01 | 0.001 | | 20.366 | -0.038 | <0.001 | 0.246 | 4.058 |
| 4 | Mother’s nervousness | 0.011 | 0.011 | 0.002 | | 28.713 | 0.028 | <0.001 | 0.247 | 4.054 |
| 5 | Exercise intensity | 0.012 | 0.012 | 0.001 | | 18.221 | -0.062 | <0.001 | 0.895 | 1.117 |
| 6 | PD^a^ | 0.013 | 0.013 | 0.001 | | 9.987 | 0.387 | 0.002 | 0.999 | 1.001 |
| Durbin-Watson | | | | | 1.96 | | | | | |
| Dependent variable: The second Principal Component | | | | | | | | | | |

a: history of physical disorder

**Table 3 Regression analysis of the third principal component on the covariates**

| **step** | **Variable** | **R2** | **Adjusted R2** | **R2 change** | | **F change** | **B** | **P** | **Tolerance** | **VIF** |
| --- | --- | --- | --- | --- | --- | --- | --- | --- | --- | --- |
| 0 | (Constant) |  |  |  | |  | -0.021 | 0.051 |  |  |
| 1 | Mother’s nervousness | 0 | 0 | 0 | | 7.36 | 0.007 | 0.007 | 1 | 1 |
| Durbin-Watson | | | | | 1.992 | | | | | |
| Dependent variable: The Third Principal Component | | | | | | | | | | |

**Table 4 Regression analysis of the fourth principal component on the covariates**

| **step** | **Variable** | **R2** | **Adjusted R2** | **R2 change** | | **F change** | **B** | **P** | **Tolerance** | **VIF** |
| --- | --- | --- | --- | --- | --- | --- | --- | --- | --- | --- |
| 0 | (Constant) |  |  |  | |  | 0.308 | <0.001 |  |  |
| 1 | Mother’s nervousness | 0.013 | 0.013 | 0.013 | | 227.33 | -0.039 | <0.001 | 0.998 | 1.002 |
| 2 | Age | 0.014 | 0.014 | 0.001 | | 19.493 | -0.012 | <0.001 | 0.999 | 1.001 |
| 3 | PCD^a^ | 0.014 | 0.014 | 0 | | 6.485 | 0.735 | 0.01 | 1 | 1 |
| 4 | Exercise intensity | 0.014 | 0.014 | 0 | | 6.109 | -0.034 | 0.013 | 0.999 | 1.001 |
| Durbin-Watson | | | | | 1.978 | | | | | |
| Dependent variable: The Fourth Principal Component | | | | | | | | | | |

a: Parent COVID-19 diagnosis status

The results of these analyses suggest that the covariates do not explain a major proportion of the residuals.
